# Job stress among workers who telecommute during the coronavirus disease (COVID-19) pandemic in Japan: a cross-sectional study

**DOI:** 10.1101/2021.03.19.21253958

**Authors:** Kazunori Ikegami, Hiroka Baba, Hajime Ando, Ayako Hino, Mayumi Tsuji, Seiichiro Tateishi, Tomohisa Nagata, Shinya Matsuda, Yoshihisa Fujino

## Abstract

**Objectives:** The work system reform and the COVID-19 pandemic in Japan have prompted efforts toward telecommuting in Japan, and there has been little research regarding the stress and health effects of telecommuting. This study aimed to clarify the relationship between telecommuting and job stress among Japanese workers.

**Study Design:** This was a cross-sectional study.

**Methods:** In December 2020, during the ‘third wave’ of the COVID-19 pandemic, an Internet-based nationwide health survey of 33,087 Japanese workers (CORoNaWork study) was conducted. Data for 27,036 individuals was included as 6,051 individuals provided invalid responses. We analysed a sample of 13,468 office workers from this database. We classified participants into four groups according to telecommuting frequency and compared the subscale of the Job Content Questionnaire and subjective job stress among these groups: high-frequency, medium-frequency, and low-frequency telecommuters group and non-telecommuters group. We used a linear mixed model and ordinal logistic regression analysis.

**Results:** There was a significant difference in the score of job control of the JCQ among the four groups after adjusting for multiple confounding factors. The high-frequency telecommuters group had the highest job control score. Regarding the fluctuation score of subjective job stress, the high- and medium-frequency telecommuters groups were significantly lower than those of the non-telecommuters group after adjusting for multiple confounding factors.

**Conclusion:** We found that high-frequency telecommuting was associated with high job control. This study suggests that the widespread adoption of telecommuting as a countermeasure to the public health challenges associated with the COVID-19 pandemic may also have a positive impact on job stress.

## Introduction

In late 2019, an outbreak of pneumonia attributed to unknown causes was confirmed in Wuhan, People’s Republic of China, and a new coronavirus (SARS-CoV-2) was discovered as the etiologic agent.^1^ Known as the coronavirus disease (COVID-19), this disease presents with various pathologies ranging from asymptomatic to severe respiratory impairment and death.^2^ The outbreak reached pandemic proportions in early 2020 and was declared a ‘Public Health Emergency of International Concern’ by the World Health Organization (WHO) in January 2020.^3^ The global COVID-19 pandemic continues to have a significant socioeconomic impact, especially in daily life, work, and medical care worldwide, including in Japan.^4-7^

In Japan, the first wave of the COVID-19 pandemic occurred in April 2020, and the second wave occurred during July–August 2020. The Japanese government declared a state of emergency in several prefectures on 7 April, 2020, subsequently expanding it nationwide on 25 May, 2020.^8^ During the declaration of the state of emergency, the people took voluntary measures to prevent infection, avoid the three Cs (crowded places, close-contact settings, confined and enclosed spaces), and not go out, with many companies implementing temporary closures and business restrictions in response to requests from local governments.^9^ In December 2020, the third wave occurred, and the number of COVID-19 infected people increased significantly compared to the previous two waves. The Japanese government declared a second state of emergency in the Tokyo metropolitan area (Tokyo, Kanagawa, Chiba, and Saitama prefectures) on 7 January, 2021, subsequently declaring it in seven prefectures (Tochigi, Aichi, Gifu, Kyoto, Osaka, Hyogo, and Fukuoka) on 13 January 2021.

The COVID-19 pandemic has had a significant influence on the work environment and work practices, and has led to changes in work system and management, such as restrictions on business trips and outings, working with physical distancing, and the digitization of customer relations.^10-13^ However, even after bringing the pandemic under control, it is speculated that some influence on people’s lives and work will continue.^14^ Particularly, telecommuting has been widely promoted as a countermeasure against emerging contagious diseases, including the COVID-19 infection.^15, 16^ The concept of telecommuting has been around since the 1970s, but current innovations in information and communication technology (ICT) have changed the work system as many workers are able to work from anywhere.^17^ In Japan, telecommuting or remote work using ICTs is being promoted to enable varied and flexible work arrangements according to individual workers’ circumstances or status through the work system reform, which became a law in 2019, and telecommuting has been strongly recommended as a countermeasure to prevent COVID-19 infection. However, since telecommuting makes it difficult to collect the information necessary for labour management, it might be difficult to check the employees’ overwork load or health problems. Furthermore, telecommuting itself might affect the physical and mental health of workers.

Several studies have investigated the health effects of telecommuting. Henke et al. reported that telecommuting might reduce health risks such as alcohol abuse, physical inactivity, tobacco use, and obesity; however, telecommuting health risks varied by telecommuting intensity, and subjects who telecommuted for 8 hours per month or less were significantly less likely to experience depression than non-telecommuters.^18^ Nijp et al. reported that working from different workplaces (such as a flexible office, home, or other remote locations) did not affect the control of working hours or the main psychosocial job factors such as job demands, job control, and social support; nevertheless, a decline in health status was observed.^19^ Several other studies on the influence of telecommuting on mental health have been reported, but there is no consensus on their views, as they may report negative or positive effects depending on various confounding factors and moderators.^20-25^

Recently, the work sysytem reform and the COVID-19 pandemic in Japan have stimulated efforts toward telecommuting in Japan, and there has been little research regarding the stress and health effects of telecommuting. This study aims to clarify the relationship between telecommuting and job stress among Japanese workers. We believe that this study provides evidence for examining the issues and coping with job stress among telecommuters during the COVID-19 pandemic.

## Methods

### Study design and setting

The Collaborative Online Research on Novel-coronavirus and Work (CORoNaWork) study is a prospective cohort study by a research group from the University of Occupational and Environmental Health. This study uses self-administered questionnaire surveys disseminated through a Japanese Internet survey company (Cross Marketing Inc. Tokyo); the baseline survey was conducted during 22– 25 December, 2020. The follow-up survey will be conducted as a cohort study with the same participants. Incidentally, during the baseline survey, the number of COVID-19 infections and deaths were overwhelmingly higher than in the first and second waves; therefore, Japan was on maximum alert during the third wave. This study adopted a cross-sectional design using a part of the data from the baseline survey of the CORoNaWork study. The details of this study protocol have been reported.^26^

### Participants

A total of 33,087 participants, who were stratified by cluster sampling by gender, age, region, and occupation, participated in the CORoNaWork study. The survey participants were between 20 and 65 years of age and were working at the time of the baseline survey. A database of 27,036 individuals was created by excluding invalid responses of 6,051 individuals. Using this database, we analysed data for 13,468 office workers. Physical workers and hospitality workers, whose jobs require mental labour, were excluded because we posited that it could be difficult for them to telecommute and may have been realistically exempted from work by telecommuting.

### Questionnaire

The questionnaire items used in this study have been described in detail.^26^ We used the data on sex, age, educational background, area of participants’ residence, job type, company size where participants work, working hours per day, family structure, telecommuting frequency, including work-related questionnaire items from the Japanese version of the Job Content Questionnaire (JCQ)^27, 28^ as well as fluctuation in subjective job stress.

The Job Content Questionnaire, developed by Karasek, is based on the job demands–control (or demand–control–support) model.^27^ The reliability and validity of the Japanese version of the JCQ has been demonstrated by Kawakami et al.^28^ We used a shortened version of the 22 items in the JCQ, in that, each item was rated on a 4-point scale (1 = strongly disagree, 4 = strongly agree). The JCQ includes a five-item job demands scales (score range: 12–48), a nine-item job control scale (score range: 24–96), a four-item supervisor support scale (score range: 4–12), and a four-item coworker support scale (score range: 4–12).

### Variables

#### Outcome variables

The scores for job demands, job control, supervisor support, and coworker support from JCQ, and the score of fluctuation in subjective job stress (1= decreased, 2=stayed the same, 3= increased) were used as outcome variables.

#### Predictor variable

We classified the participants into four groups according to telecommuting frequency: high-frequency telecommuters group for participants telecommuting for four days or more per week; medium-frequency, for those telecommuting for two to three days a week; low-frequency, for those telecommuting for one day or less in a week; and non-telecommuters group, for those not telecommuting. These variables were used as the predictor variables.

#### Potential confounders

The following items, surveyed using a questionnaire, were used as confounding factors. Sex, age, and education (junior or senior high school, junior college or vocational school, university, or graduate school) were the personal characteristics. Occupation (regular employees, managers, executives, public service worker, temporary workers, freelancers or professionals, others), company size where participants work (≤9, 10-49, 50-99, 100-499, 500-999, 1000-9999, ≥10000 employees), working hours per day (<8, ≥8 and < 9, ≥9 and <11, ≥11) were used as work-related factors, while marital status (married, unmarried) and living with family (presence or absence) were the familial factors. Additionally, the prefecture of participants’ residence was used as another variable.

### Statistical analyses

We used a linear mixed model (LMM) to analyse the relationships between the four groups of telecommuting frequency and the subscales of the JCQ. In this stage, the dependent variables consisted of the scores of job demand, job control, coworker support, and supervisor support of JCQ, and the following three models were analysed. In Model 1, we treated the four classifications of telecommuting frequency as fixed effects and treated the prefecture of residence as random effects. In Model 2, we added the personal characteristics variables to the fixed effects of Model 1. In Model 3, we added the work-related and familial variables to the fixed effects of Model 2. The estimated marginal means (EMM) of the subscale by four groups of telecommuting frequency was calculated by adjusting for the dependent variable in each model of LMM. Akaike’s information criterion (AIC) was used to determine the goodness of fit of the statistical model.

We employed an ordinal logistic regression analysis (OLR) to analyse four classifications of telecommuting frequency and the fluctuation in subjective job stress. The dependent variable consisted of the fluctuation in subjective job stress, and the following three models were analysed. In Model 1, as a crude analysis, we treated the four classifications of telecommuting frequency as the independent variable. In Model 2, we adjusted for the personal characteristics variables. In Model 3, we adjusted for personal characteristics, work-related factors, and familial factors. Cox and Snell R-squared was used to determine the goodness of fit of the statistical model. In all tests, the threshold for significance was set at p<0.05. SPSS 25.0J analytical software (IBM, NY) was used for the statistical analyses.

## Results

### Participants and descriptive data

There were 2,042 participants in the high-frequency telecommuters group, 1,058 in the medium-frequency telecommuters group, 952 in the low-frequency telecommuters group, and 9,416 in the non-telecommuters group (Fig. 1). The participant characteristics classified by the telecommuting frequency groups are shown in Table 1. Male participants telecommuted more than female participants. The workers aged 50 and over tended to telecommute often. Regarding work-related factors, the proportion of those who belonged to companies with ≤9 employees and those who worked less than 8 hours/day was high in the high-frequency telecommuters group. Regarding familial factors, the proportion of those who were married and were living together with their family was low in the high-frequency telecommuters group (Table 1).

**Fig. 1.**
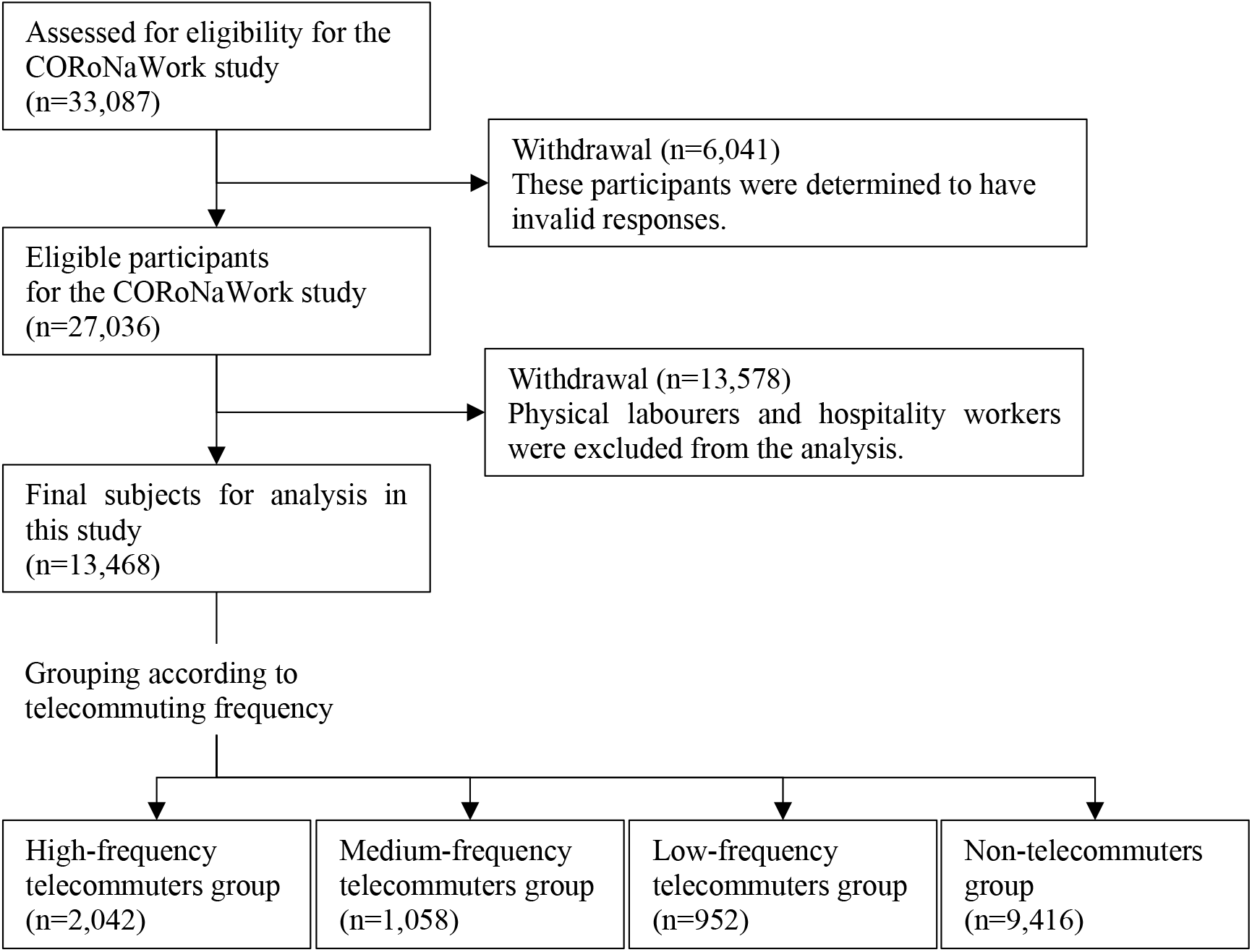
Flow chart of this study population selection.

**Table 1.** Participants’ characteristics by telecommuting frequency groups; n (%)

| Items | Total |  | Telecommuting frequency |  |  |  |  |  |  |  |
| --- | --- | --- | --- | --- | --- | --- | --- | --- | --- | --- |
|  | (n=13468) |  | High (n=2042) |  | Medium (n=1058) |  | Low (n=952) |  | None (n=9416) |  |
|  | n | (%) | n | (%) | n | (%) | n | (%) | n | (%) |
| Sex, male | 6896 | (51.2) | 1159 | (56.8) | 591 | (55.9) | 613 | (64.4) | 4533 | (48.1) |
| Age (in years) |  |  |  |  |  |  |  |  |  |  |
| 20-29 | 751 | (5.6) | 81 | (4.0) | 52 | (4.9) | 54 | (5.7) | 564 | (6.0) |
| 30-39 | 2227 | (16.5) | 308 | (15.1) | 170 | (16.1) | 142 | (14.9) | 1607 | (17.1) |
| 40-49 | 4080 | (30.3) | 558 | (27.3) | 292 | (27.6) | 278 | (29.2) | 2952 | (31.4) |
| 50-59 | 4682 | (34.8) | 765 | (37.5) | 383 | (36.2) | 345 | (36.2) | 3189 | (33.9) |
| ≥60 | 1728 | (12.8) | 330 | (16.2) | 161 | (15.2) | 133 | (14.0) | 1104 | (11.7) |
| Education |  |  |  |  |  |  |  |  |  |  |
| Junior or senior high school | 3018 | (22.4) | 366 | (17.9) | 119 | (11.2) | 132 | (13.9) | 2401 | (25.5) |
| Junior college or vocational school | 2780 | (20.6) | 433 | (21.2) | 163 | (15.4) | 138 | (14.5) | 2046 | (21.7) |
| University or graduate school | 7670 | (56.9) | 1243 | (60.9) | 776 | (73.3) | 682 | (71.6) | 4969 | (52.8) |
| Occupation |  |  |  |  |  |  |  |  |  |  |
| Regular employees | 6483 | (48.1) | 647 | (31.7) | 522 | (49.3) | 434 | (45.6) | 4880 | (51.8) |
| Managers | 1743 | (12.9) | 173 | (8.5) | 199 | (18.8) | 225 | (23.6) | 1146 | (12.2) |
| Executives | 545 | (4.0) | 96 | (4.7) | 53 | (5.0) | 52 | (5.5) | 344 | (3.7) |
| Public service worker | 1758 | (13.1) | 25 | (1.2) | 58 | (5.5) | 107 | (11.2) | 1568 | (16.7) |
| Temporary workers | 1411 | (10.5) | 157 | (7.7) | 108 | (10.2) | 75 | (7.9) | 1071 | (11.4) |
| Freelancers or professionals | 1257 | (9.3) | 756 | (37.0) | 96 | (9.1) | 45 | (4.7) | 360 | (3.8) |
| Others | 271 | (2.0) | 188 | (9.2) | 22 | (2.1) | 14 | (1.5) | 47 | (0.5) |
| Company size |  |  |  |  |  |  |  |  |  |  |
| ≤9 employees | 2732 | (20.3) | 1100 | (53.9) | 174 | (16.4) | 110 | (11.6) | 1348 | (14.3) |
| 10-49 employees | 2009 | (14.9) | 109 | (5.3) | 98 | (9.3) | 90 | (9.5) | 1712 | (18.2) |
| 50-99 employees | 1164 | (8.6) | 69 | (3.4) | 70 | (6.6) | 65 | (6.8) | 960 | (10.2) |
| 100-499 employees | 2543 | (18.9) | 160 | (7.8) | 167 | (15.8) | 164 | (17.2) | 2052 | (21.8) |
| 500-999 employees | 1038 | (7.7) | 98 | (4.8) | 94 | (8.9) | 96 | (10.1) | 750 | (8.0) |
| 1000-9999 employees | 2767 | (20.5) | 306 | (15.0) | 274 | (25.9) | 263 | (27.6) | 1924 | (20.4) |
| ≥10000 employees | 1215 | (9.0) | 200 | (9.8) | 181 | (17.1) | 164 | (17.2) | 670 | (7.1) |
| Working hours per day (h/d) |  |  |  |  |  |  |  |  |  |  |
| < 8h/d | 2658 | (19.7) | 723 | (35.4) | 233 | (22.0) | 164 | (17.2) | 1538 | (16.3) |
| 8≤ and <9h/d | 7769 | (57.7) | 904 | (44.3) | 558 | (52.7) | 509 | (53.5) | 5798 | (61.6) |
| 9≤ and <11h/d | 2583 | (19.2) | 341 | (16.7) | 237 | (22.4) | 241 | (25.3) | 1764 | (18.7) |
| ≥11h/d | 458 | (3.4) | 74 | (3.6) | 30 | (2.8) | 38 | (4.0) | 316 | (3.4) |
| Marriage status, married | 7764 | (57.6) | 991 | (48.5) | 650 | (61.4) | 625 | (65.7) | 5498 | (58.4) |
| Presence of family living together | 10642 | (79.0) | 1512 | (74.0) | 828 | (78.3) | 770 | (80.9) | 7532 | (80.0) |

The groups according to telecommuting frequency are High: high-frequency telecommuters group, Medium: medium-frequency telecommuters group, Low; low-frequency telecommuters group, None; non-telecommuters group.

### The subscales of the JCQ among the telecommuting frequency groups

We compared the scores of each subscale of the JCQ among the four groups of telecommuting frequency using LMM (models 1–3) (Table 2).

**Table 2.** Comparison of the scores of subscales of the JCQ among the telecommuting frequency groups

| Parameters | Telecommuting frequency | Model 1 |  |  | Model 2 |  |  | Model 3 |  |  |
| --- | --- | --- | --- | --- | --- | --- | --- | --- | --- | --- |
|  |  | EMM | 95%CI | p | EMM | 95%CI | p | EMM | 95%CI | p |
| Job demands | High | 28.3 | 28.0-28.5 | <0.001 | 28.1 | 27.8-28.4 | <0.001 | 29.9 | 29.6-30.2 | 0.332 |
|  | Medium | 29.3 | 28.9-29.7 |  | 29.0 | 28.7-29.4 |  | 30.0 | 29.6-30.4 |  |
|  | Low | 29.6 | 29.2-30.0 |  | 29.2 | 28.8-29.6 |  | 30.0 | 29.6-30.4 |  |
|  | None | 29.4 | 29.3-29.5 |  | 29.2 | 29.1-29.4 |  | 30.2 | 29.9-30.4 |  |
| Job control | High | 68.6 | 68.1-69.1 | <0.001 | 67.6 | 67.1-68.2 | <0.001 | 68.3 | 67.7-69 | <0.001 |
|  | Medium | 66.2 | 65.4-66.9 |  | 65.2 | 64.5-65.9 |  | 67.2 | 66.4-68 |  |
|  | Low | 66.2 | 65.5-66.9 |  | 65.0 | 64.2-65.7 |  | 67.0 | 66.2-67.8 |  |
|  | None | 62.6 | 62.3-62.9 |  | 62.2 | 61.8-62.5 |  | 64.7 | 64.2-65.2 |  |
| Supervisor support | High | 9.8 | 9.7-10.0 | <0.001 | 9.8 | 9.6-9.9 | <0.001 | 9.9 | 9.7-10.1 | 0.011 |
|  | Medium | 10.3 | 10.2-10.5 |  | 10.2 | 10-10.4 |  | 10.0 | 9.8-10.2 |  |
|  | Low | 10.4 | 10.3-10.6 |  | 10.3 | 10.1-10.5 |  | 10.0 | 9.8-10.2 |  |
|  | None | 10.0 | 10.0-10.1 |  | 10.0 | 9.9-10.1 |  | 9.8 | 9.6-9.9 |  |
| Coworker support | High | 10.1 | 10.0-10.2 | <0.001 | 10.1 | 10-10.2 | <0.001 | 10.2 | 10-10.3 | 0.051 |
|  | Medium | 10.6 | 10.4-10.7 |  | 10.5 | 10.4-10.7 |  | 10.3 | 10.1-10.5 |  |
|  | Low | 10.7 | 10.6-10.9 |  | 10.7 | 10.5-10.9 |  | 10.4 | 10.2-10.6 |  |
|  | None | 10.4 | 10.4-10.5 |  | 10.4 | 10.4-10.5 |  | 10.2 | 10.1-10.3 |  |

EMM: Estimated marginal mean, CI: Confidence interval. The groups according to telecommuting frequency are High (H): high-frequency telecommuters group, Medium (M): medium-frequency telecommuters group, Low (L); low-frequency telecommuters group, None (N); non-telecommuters group.

In each of the four subscales of the JCQ, the fit of the statistical model as determined by AIC was the best for Model 3 and the worst for Model 1. There were significant differences in the score of job demands among the four groups in Models 1 and 2, and the high-frequency telecommuters group had the lowest score of job demands. However, there was no significant difference in the score of job demands among the four groups in Model 3. There was a significant difference in the score of job control among the four groups in all the models. The high-frequency telecommuters group had the highest score of job control, while that of the non-telecommuters groups was the lowest. There were significant differences in the score of supervisor support among the four groups in all the models. The high-frequency telecommuters group had the lowest score of supervisor support in Models 1 and 2, but not in Model 3. In Model 3, the non-telecommuters group had the lowest score of supervisor support. There were significant differences in the score of coworker support among the four groups in Models 1 and 2, and the coworker support of high-frequency telecommuters group had the lowest scores in Model 1 and 2. However, there were no significant differences among the four groups in Model 3.

### The score of fluctuation in subjective job stress among the telecommuting frequency groups

The distribution of the score (1-3) of subjective job stress classified by telecommuting frequency is shown in Table 3. We compared the scores of fluctuation in subjective job stress among the four groups using OLR (Models 1–3) (Table 4). The fit of a statistical model as determined by Cox and Snell R-squared model was the best for Model 3 and the worst for Model 1. In all models, the subjective job stress scores of the high- and medium-frequency telecommuters groups were significantly lower than that of the non-telecommuters group. (Reference).

**Table 3.**
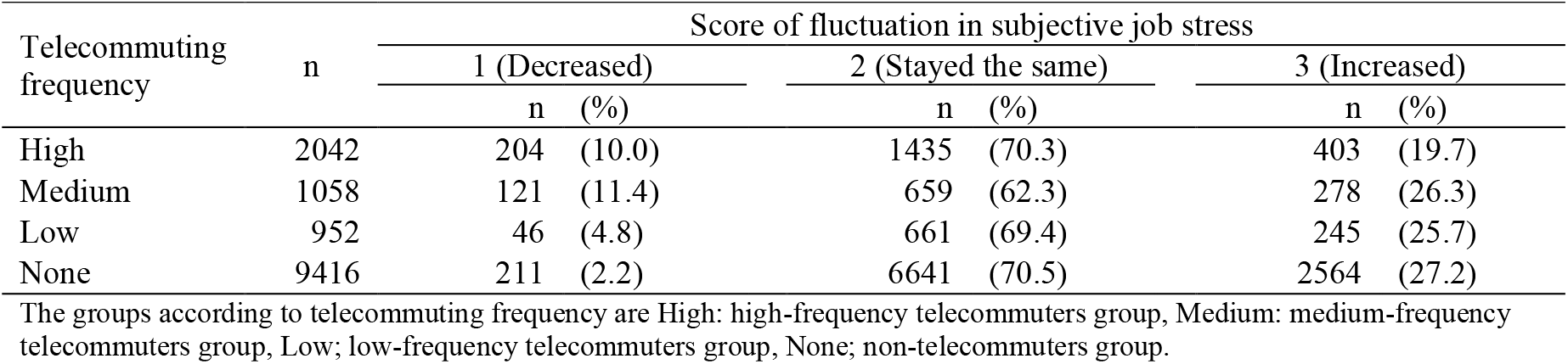
Distribution of the score of fluctuation in subjective job stress according to telecommuting frequency groups

| Telecommuting frequency | n | Score of fluctuation in subjective job stress |  |  |  |  |  |
| --- | --- | --- | --- | --- | --- | --- | --- |
|  |  | 1 (Decreased) |  | 2 (Stayed the same) |  | 3 (Increased) |  |
|  |  | n | (%) | n | (%) | n | (%) |
| High | 2042 | 204 | (10.0) | 1435 | (70.3) | 403 | (19.7) |
| Medium | 1058 | 121 | (11.4) | 659 | (62.3) | 278 | (26.3) |
| Low | 952 | 46 | (4.8) | 661 | (69.4) | 245 | (25.7) |
| None | 9416 | 211 | (2.2) | 6641 | (70.5) | 2564 | (27.2) |

The groups according to telecommuting frequency are High: high-frequency telecommuters group, Medium: medium-frequency telecommuters group, Low; low-frequency telecommuters group, None; non-telecommuters group.

**Table 4.** Comparison of the fluctuation score of subjective job stress among telecommuting frequency groups

| Telecommuting frequency | Model 1 |  |  | Model 2 |  |  | Model 3 |  |  |
| --- | --- | --- | --- | --- | --- | --- | --- | --- | --- |
|  | OR | 95%CI | p | OR | 95%CI | p | OR | 95%CI | p |
| High | 0.52 | 0.46-0.58 | <0.001 | 0.54 | 0.48-0.6 | <0.001 | 0.54 | 0.47-0.61 | <0.001 |
| Medium | 0.68 | 0.59-0.79 | <0.001 | 0.71 | 0.62-0.82 | <0.001 | 0.72 | 0.63-0.84 | <0.001 |
| Low | 0.86 | 0.74-0.99 | 0.038 | 0.90 | 0.77-1.04 | 0.143 | 0.90 | 0.78-1.04 | 0.164 |
| None | reference |  |  | reference |  |  | reference |  |  |

OR: Odds ratio, CI: Confidence interval. The groups according to telecommuting frequency are High: high-frequency telecommuters group, Medium: medium-frequency telecommuters group, Low; low-frequency telecommuters group, None; non-telecommuters group.

## Discussion

This study clarified the relationship between job stress and telecommuting frequency. First, we consider the relationships between job demands and control and telecommuting frequency. When adjusted only for residence and personal characteristics, high-frequency telecommuters had significantly lower job demands than others. However, after adjusting for work-related and familial factors, we observed no significant difference between job demands and telecommuting frequency. The high-frequency telecommuters group had a higher proportion of participants who worked less than eight hours per day. Therefore, it is reasonable to assume that job demands could be more influenced by working hours than telecommuting frequency. Job control was higher for high-frequency telecommuters, even after adjusting for residence, personal characteristics, and work-related and familial factors. We believe that this finding is important. Previously, it was supposed that most workers who can telecommute could be employed in specific job positions or occupations where they can decide their work contents or could have the authority to determine their own work hours (such as flexible work hours). On the other hand, to control the current COVID-19 pandemic, many workers may be forced to telecommute at the behest of the Japanese government. Despite the ad hoc telecommuting measures during the COVID-19 pandemic, it is possible that it has had a positive influence. These might lead to improved job control and reduced job stress by preparing for appropriate situations where workers can telecommute.

Next, we consider the relationship between social support and telecommuting. When adjusted for residence and personal characteristics, high-frequency telecommuters had significantly lower supervisor and coworker support than others. However, when adjusted for work-related and familial factors, we could not observe any significant differences by grouping the telecommuting frequency. In this regard, we believe that the influence of familial factors could be important. In this study, the proportion of participants who were unmarried and not living with family is high in the high-frequency telecommuters group. Therefore, these familial factors might lead to poor communication and reduced social support scores. The psychological repercussions of balancing work and family life has long been proposed in the concept of Work–Family Conflict.^29^ The relationship between work and family has important effects on job and life satisfaction; especially, an appropriate assignment between an individual’s work and family roles increases job and family satisfaction.^30^ Furthermore, telecommuting has been reported to lead to work flexibility and increased time spent at home, which increased happiness and life satisfaction and decreased stress associated with work–life balance.^31^ In this context, telecommuting may have indirectly influenced the coping behaviour associated with Work-Family Conflict^32^ and complementarily influenced the social support at work.

When adjusted for residence, personal characteristics, and work-related and familial factors, the score of fluctuation in subjective job stress of high-frequency telecommuters and medium-frequency telecommuters was significantly lower than that of non-telecommuters. One of the reasons could be that high job control reduces subjective job stress. However, job stress is influenced by various factors; particularly, the impact of the COVID-19 pandemic must also be considered. With the COVID-19 pandemic, many people are worried about infection in their daily lives and also in the workplace. Telecommuting is an effective measure for preventing COVD-19 infection, and it may have had a positive effect on reducing job stress such as anxiety and depression.^33 34^ We speculate that the elimination of infection anxiety related to COVID-19 has influenced the reduction in work stress.

### Limitations

This study has four limitations. First, since the CORoNaWork study is an Internet-based survey, the generalisability of the results is uncertain. However, we attempted to reduce the bias of the subjects as much as possible by sampling them by generation, residence, and occupation. Second, since the present study adopted a cross-sectional design, the causal relationship between telecommuting and job stress is unknown. Telecommuting has been strongly recommended from the early phase of COVID-19 pandemic as one of the measures to prevent the spread of COVID-19 infection in the workplace,^8^ and it continues to be in force now. We conducted this study more than half a year since the start of the COVID-19 pandemic, and thus, we believe that the context of the impact of telecommuting on job stress is reasonable. Third, regarding the fluctuation in subjective job stress, we assessed the degree of fluctuation from the past to the present using a three-point Likert scale. Therefore, the existence of a response bias cannot be denied. It is necessary to reconsider the questionnaire items and conduct a longitudinal evaluation. Fourth, because this study assessed telecommuting during the COVID-19 pandemic, there may be some differences in the effects on job stress during normal telecommuting. However, this study was consistent with previous studies and adjusted for potential confounders to ensure a certain degree of rationality. Further research on telecommuting in Japan is required.

## Conclusion

In this study, we analysed the relationship between job stress and telecommuting in Japanese workers during the COVID-19 pandemic using the CORoNaWork database. We found that high-frequency telecommuting was associated with high job control among the four job stressors such as job demands, job control, supervisor support, and coworker support. Additionally, telecommuting might reduce subjective job stress, but this may have limited influence in the context of the COVID-19 pandemic.

## Data Availability

Due to the nature of this research, the participants of this study did not agree to their data being shared publicly, and hence, supporting data is not available.

## Declaration

### Notes

This article has submitted for peer-review and publication in “International Journal of Occupational Medicine and Environmental Health” (http://ijomeh.eu).

## Acknowledgements

We appreciate all the participants and all members of the CORoNaWork Study Group. The current members of the CORoNaWork Project, in alphabetical order, are as follows: Dr. Yoshihisa Fujino (present chairperson of the study group), Dr. Akira Ogami, Dr. Arisa Harada, Dr. Ayako Hino, Dr. Hajime Ando, Dr. Hisashi Eguchi, Dr. Kazunori Ikegami, Dr. Kei Tokutsu, Dr. Keiji Muramatsu, Dr. Koji Mori, Dr. Kosuke Mafune, Dr. Kyoko Kitagawa, Dr. Masako Nagata, Dr. Mayumi Tsuji, Ms. Ning Liu, Dr. Rie Tanaka, Dr. Ryutaro Matsugaki, Dr. Seiichiro Tateishi, Dr. Shinya Matsuda, Dr. Tomohiro Ishimaru, and Dr. Tomohisa Nagata. All members are affiliated with the University of Occupational and Environmental Health, Japan. We would also like to thank Editage (www.editage.com) for English language editing.

## Author contributions

KI; writing the manuscript and analysis, HB, HA, AH; analysis, review of manuscripts, and advice on interpretation AH; review of manuscripts, and advice on interpretation, TN; review of manuscripts and advice on interpretation, ST, MT, and SM; review of manuscripts, advice on interpretation, and funding for research, YF; overall survey planning, creating the questionnaire, analysis, and drafting the manuscript, all authors have read and agreed to the published version of the manuscript.

## Ethical approval

This study was approved by the Ethics Committee of the University of Occupational and Environmental Health, Japan.

